# Travel-time, bikes, and HIV elimination in Malawi: a geospatial modeling analysis

**DOI:** 10.1101/2020.09.01.20186072

**Authors:** Laurence Palk, Justin T. Okano, Luckson Dullie, Sally Blower

## Abstract

**Background:** UNAIDS has prioritized Malawi and 21 other countries in sub-Saharan Africa (SSA) for “fast-tracking” the end of their HIV epidemics. To achieve elimination requires treating 90% of people living with HIV (PLHIV); coverage is already fairly high (70–75%). However, many individuals in SSA have to walk to access healthcare. We use data-based geospatial modeling to determine whether the need to travel long distances to access treatment and limited transportation in rural areas are barriers to HIV elimination in Malawi. Additionally, we evaluate the effect on treatment coverage of increasing the availability of bicycles in rural areas.

**Methods:** We build a geospatial model that we use to estimate, for every PLHIV, their travel-time to access HIV treatment if driving, bicycling, or walking. We estimate the travel-times needed to achieve 70% or 90% coverage. Our model includes a spatial map of healthcare facilities (HCFs), the geographic coordinates of residencies for all PLHIV, and an “impedance” map. We quantify impedance using data on road/river networks, land cover, and topography.

**Findings:** To cross an area of one km^2^ in Malawi takes from ~60 seconds (driving on main roads) to ~60 minutes (walking in mountainous areas); ~80% of PLHIV live in rural areas. At ~70% coverage, HCFs can be reached within: ~45 minutes if driving, ~65 minutes if bicycling, and ~85 minutes if walking. Increasing coverage above ~70% will become progressively more difficult. To achieve 90% coverage, the travel-time for many PLHIV (who have yet to initiate treatment) will be almost twice as long as those currently on treatment. Increasing bicycle availability in rural areas reduces round-trip travel-times by almost one hour (in comparison with walking), and could substantially increase coverage levels.

**Interpretation:** Geographic inaccessibility to treatment coupled with limited transportation in rural areas are substantial barriers to reaching 90% coverage in Malawi. Increased bicycle availability could help eliminate HIV.

**Funding:** National Institute of Allergy and Infectious Diseases

## Introduction

UNAIDS has proposed an ambitious strategy for ending the HIV pandemic by 2030.^1^ For this strategy to succeed, each HIV-afflicted country needs to treat 90% of people living with HIV infection (PLHIV).^1^ Governments in sub-Saharan Africa (SSA), where ~26 million people are PLHIV, are striving to reach this 90% coverage goal. UNAIDS has prioritized 22 countries in SSA for “fast-tracking” the end of their HIV epidemic.^1^ Several of these countries (e.g., Malawi, Lesotho, Zimbabwe) have already reached fairly high coverage levels (70%–75%).^2-4^ Finding and treating the last 25–30% of PLHIV, many of whom live in rural areas, is likely to be extremely challenging. Throughout SSA, distance to healthcare facilities (HCFs) and lack of transportation are major obstacles to accessing HIV treatment.^5-11^ Here we use data-based geospatial modeling to evaluate their potential impact as barriers to HIV elimination in Malawi, one of UNAIDS’ designated Fast-Track countries.^12^ Specifically, we quantify their impact on preventing treatment coverage from reaching UNAIDS’ target of 90%. In Malawi, as in other sub-Saharan African countries, many PLHIV have to walk to access treatment.^8,9^ Therefore, we also use our modeling framework to evaluate the potential impact of a novel intervention strategy for improving treatment coverage in Malawi: increasing the availability of bicycles in rural areas. We discuss the policy implications of our results for Malawi and other HIV-afflicted countries in SSA.

Malawi is one of the most severely HIV-afflicted countries in SSA: 9–10% of the population are PLHIV.^2,13^ Malawi is also one of the poorest country in the world; ~40% of the population of ~18.6 million^14^ live in poverty. The population is predominantly (83%) rural^15^, there are only four cities in the country (figure 1A). Population density varies from extremely low in some rural areas, less than 10 individuals per km^2^, to more than 10,000 individuals per km^2^ in cities (figure S1; appendix p 6).^16^ In the rural areas, transportation is very limited: ~2% of households have a car or truck and less than 10% of individuals own a bicycle.^13^ Furthermore, not all of these bicycles are functional; many are in a poor state of repair. Notably, in some rural areas, patients, such as pregnant women, are transported to the nearest HCF by a bicycle ambulance.

**Figure 1:**
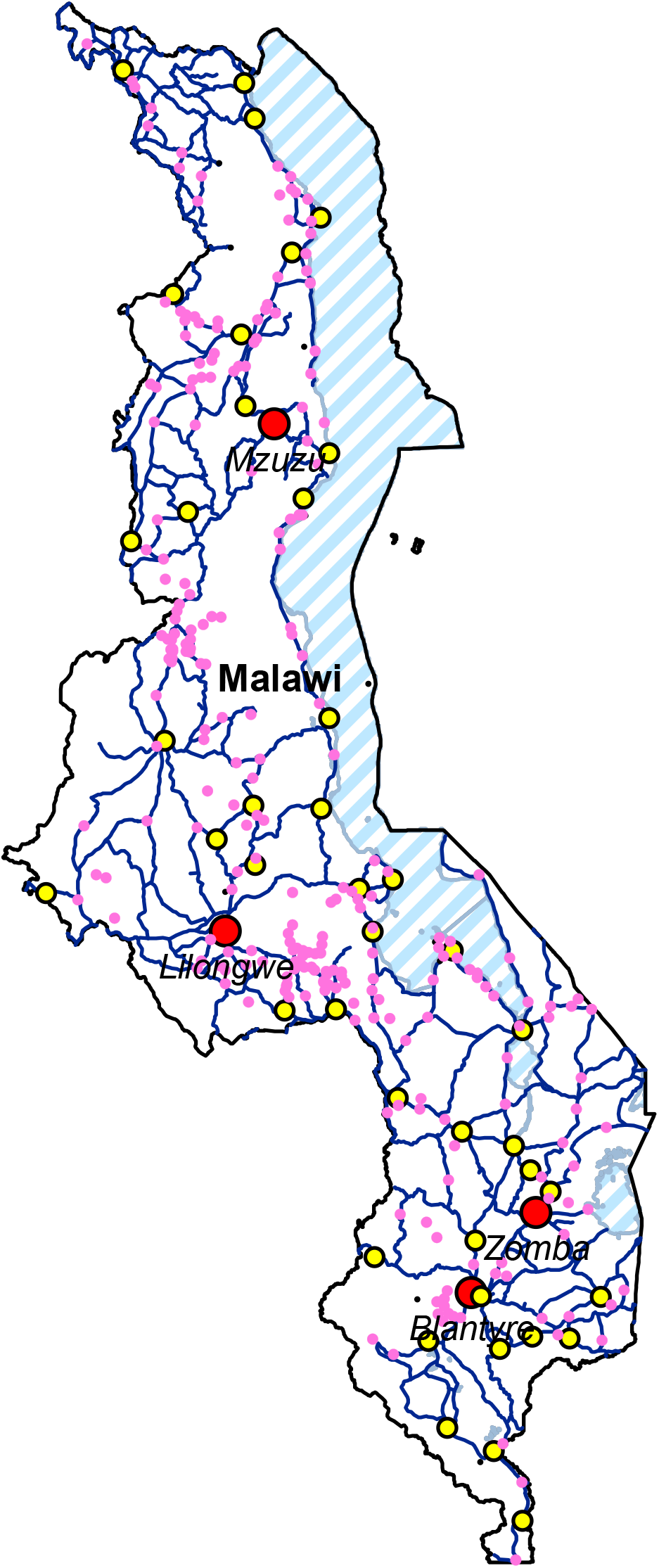
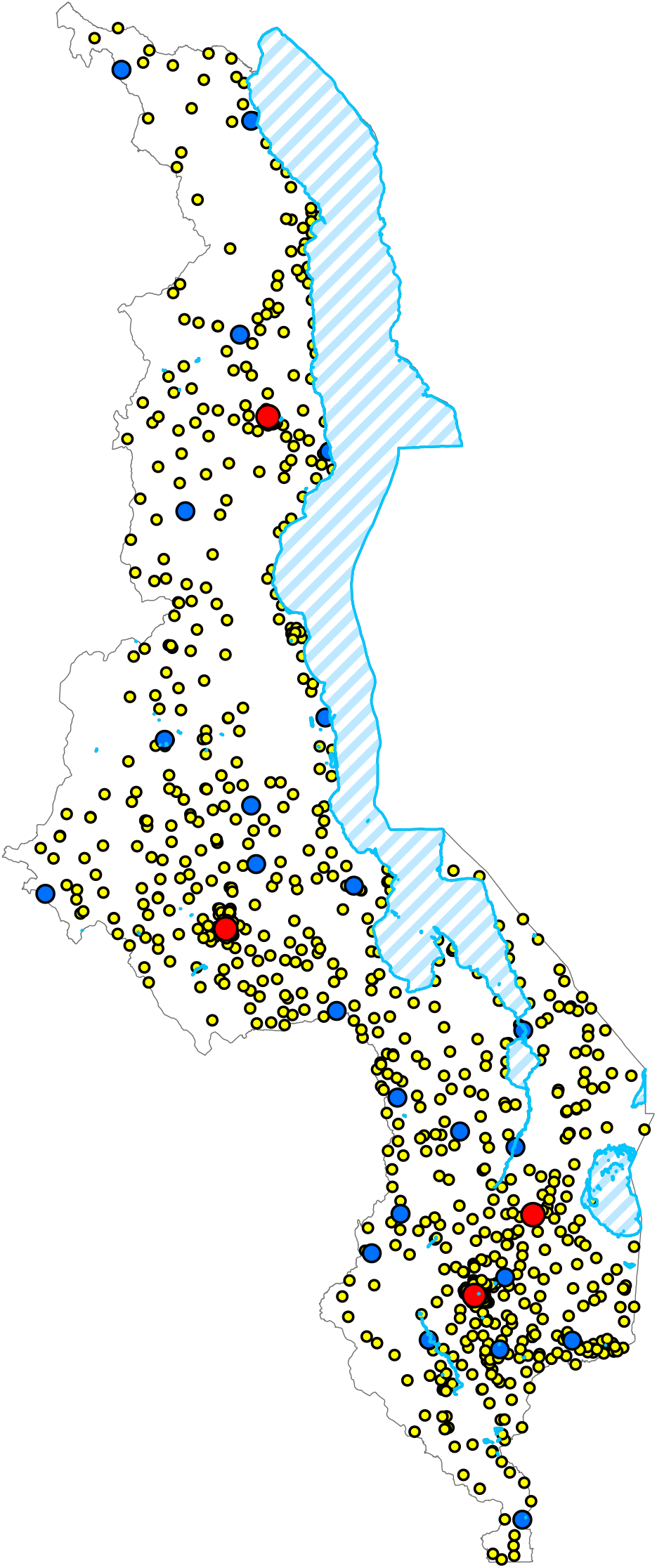
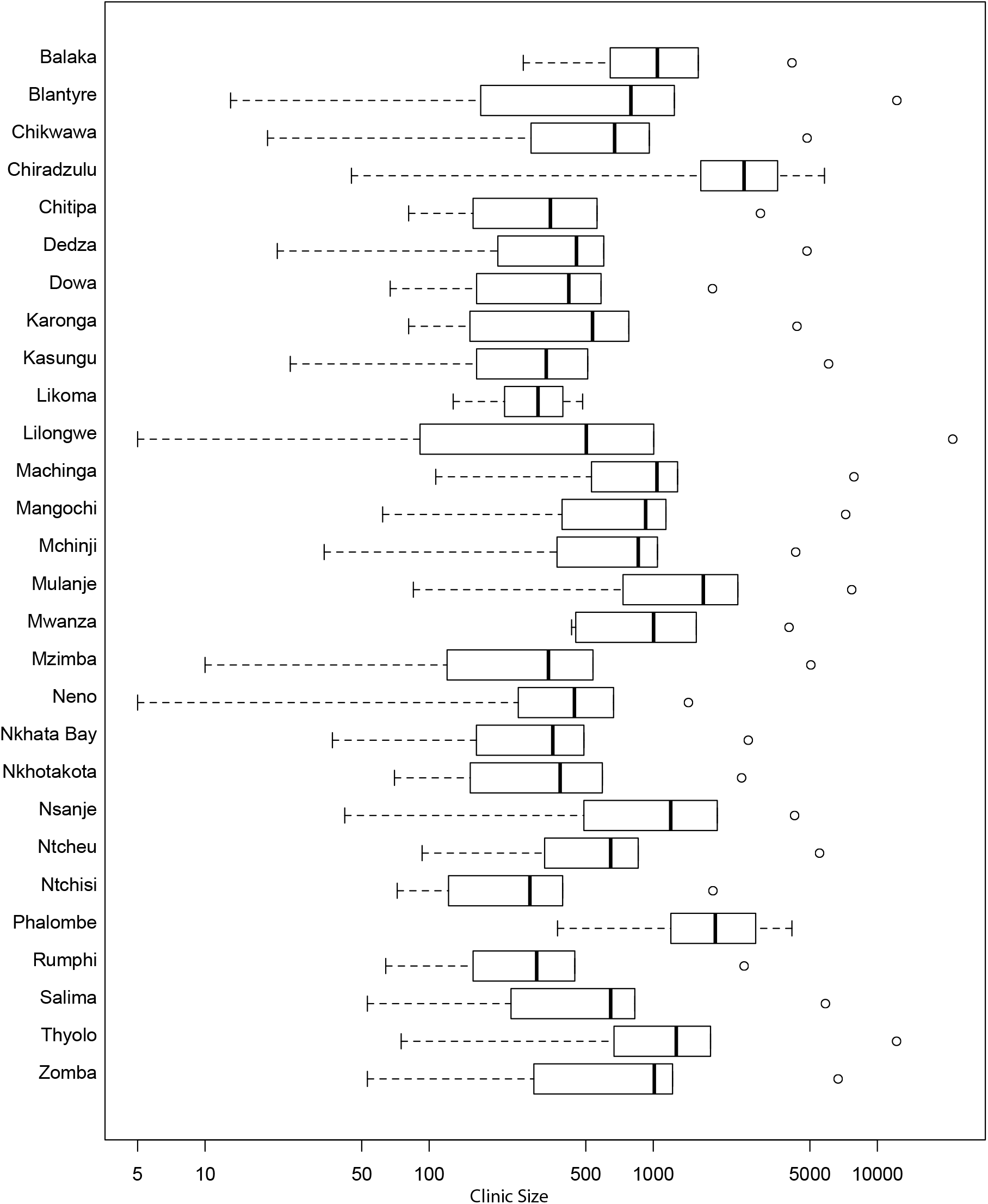
Geography of Malawi and healthcare infrastructure. **(A)** Map of Malawi. Striped blue areas represent lakes, red circles cities, yellow circles towns, pink dots villages, and blue lines roads. There are only four cities in Malawi: Mzuzu in the north, Lilongwe (the capital) in the central region, and Blantyre and Zomba in the south. (**B)** Map of Malawi’s health care facilities: central hospitals (red circles), district hospitals (blue circles), and small hospitals and clinics (yellow circles). **(C)** Boxplots show the numbers of patients registered as alive and receiving HIV treatment at every HCF in each of the 28 healthcare districts in Malawi. Clinic size is plotted on a logarithmic scale.

The Government of Malawi began the rollout of HIV treatment in 2004,^17^ and made it available for free.^18^ The focus of the Governments’ current National Strategic Plan (NSP) for HIV is to have reached UNAIDS’ 2020 goals by the end of 2020.^17^ Specifically, they aim to have diagnosed 90% of all PLHIV, initiated and retained 90% of those diagnosed on treatment, and achieved viral suppression for 90% of those on treatment. Currently, 77% of PLHIV know their status, 91% are on treatment and 91% have viral suppression.^2^ As a consequence, the current coverage of treatment is 70%.^2^ The NSP is heavily focused on HIV testing as the gateway to reaching the 90 90 90 targets. Malawi has been shifting from the community-based testing model, which focuses on providing access to the general population, to a more targeted approach on key and vulnerable populations. Key populations include female sex workers and men who have sex with men; vulnerable populations include prisoners, youth, and highly mobile groups. These populations are those in which the incidence and prevalence of HIV is the highest. This targeted approach is being taken in order to maximize the efficiency of testing and realize the highest yield; i.e., identifying the maximum number of undiagnosed PLHIV per dollar spent. After reaching UNAIDS 2020 90 90 90 goals, Malawi aims to achieve UNAIDS’ 2030 95 95 95 goals.

Malawi’s healthcare system is decentralized; it is organized at four levels: community, primary, secondary and tertiary.^19,20^ These different levels are linked to each other through a referral system. At the community level, health services are provided by health posts, dispensaries, village clinics, maternity clinics, mobile clinics, and health surveillance assistants. Each assistant is responsible for a catchment area of 1,000 individuals. At the primary level, health services are provided by health centers and community hospitals. Health centers offer outpatient and maternity services and are meant to serve a population of 10,000. Community hospitals are larger than health centers. They offer outpatient and inpatient services and conduct minor procedures. Their bed capacity can reach up to 250 beds. At the community and primary level, together, there are ~950 clinics and small hospitals. The secondary level of care consists of 28 district hospitals. They provide referral services to health centers and community hospitals and also provide their surrounding populations with both outpatient and inpatient services. The tertiary level consists of four central hospitals. They provide specialist health services at the regional level and also referral services to district hospitals within their region.

Malawi now has 750 HCFs that provide HIV-testing and treatment throughout the country. These include both public and private facilities. The geographic distribution of Malawi’s healthcare system is shown in figure 1B. Many of the HCFs in the rural areas are extremely small with some treating as few as five PLHIV; the largest HCF, which is in the capital city (Lilongwe), provides treatment to ~20,000 PLHIV (figure 1C). Recently, UNICEF estimated that 45% of people in Malawi live more than five kilometers from a HCF.^21^ Malawi’s Ministry of Health, in order to remedy this situation, is evaluating the financial expenditure that would be required to build 900 new health posts in rural areas.

To conduct our analyses, we built a geospatial modeling framework that enabled us to estimate - for every PLHIV in Malawi - how long it would take them to travel from where they live to their nearest HCF. We define this length of time as the travel-time to treatment. We estimated travel-time to HCFs rather than simply estimating the distance to HCFs, because travel-time (which incorporates distance) provides a more realistic measure of geographic accessibility. We calculated travel-times based on three modes of transportation: driving, bicycling, or walking. To calculate travel-times we needed to: (i) know the geographic coordinates of all of the HCFs in Malawi, (ii) estimate the geographic coordinates of the residencies of every PLHIV in Malawi, and (iii) quantify the “impedance” to travelling across Malawi. We obtained the coordinates of the HCFs from the Ministry of Health in Malawi,^19^ and estimated the coordinates of the residencies of all PLHIV by constructing a Density of Infection (DoI) map.^22,23^ We defined impedance in terms of the time it would take an (average) individual to travel through an area of one square kilometer, anywhere in Malawi, when using a specific mode of transportation.^24^ To quantify impedance, we used data on road and water body networks throughout the country, the type of roads, the type of land cover, and the topography of the landscape.

## Methods

Our geospatial modeling framework has three components: a spatial map of the healthcare infrastructure in Malawi (figure 1B), a country-level DoI map^22,23^ and a Friction Surface Raster map.^24^ The Friction Surface Raster map enabled us to quantify impedance to travel throughout Malawi.

We constructed the DoI map by using raster multiplication to combine gridded WorldPop data on population density and settlement patterns^16^ with an Epidemic Surface Prevalence map.^22^ The WorldPop data have been mapped to a spatial resolution of 100 meters by 100 meters;^16^ the accuracy of these projections has been assessed elsewhere.^25^ The Epidemic Surface Prevalence map was constructed by spatially smoothing HIV-testing data collected from ~16,000 adults (15–49 years old) who participated in the 2015 Malawi Demographic and Health Survey (MDHS).^13^ Detailed methods for constructing the Epidemic Surface Prevalence map and the DoI map are given in the appendix (pp 2–3). The appendix (pp 3–4) also describes the methodology for, and results from, an uncertainty and sensitivity analysis.

The MDHS is a nationally representative sample of Malawi’s population, collected using a two-stage cluster design. Figure S2 (appendix p 7) shows the location of the clusters: 173 were urban, 677 were rural. Demographic, socio-economic, behavioral, and biometric data were collected; blood samples were drawn from women aged 15–49 and men aged 15–54 to determine their HIV infection status. All women 15–49 and men 15–54 who were permanent residents of sampled households (or visitors who had stayed over the night prior to the survey) were eligible for HIV-testing. Participation in HIV-testing was 93% for women aged 15–49 and 87% for men aged 15–54. Participation rates for HIV-testing are for individuals who were at home at the time of the survey. Testing data were georeferenced using the location of the sampling cluster.

We constructed the Friction Surface Raster map by using AccessMod software^24^ to combine raster images of road and water body networks (figure 2A) with topography (figure 2B) and land cover (figure 2C). The software includes an algorithm that calculates travel speeds based on land cover, the type of road, and mode of transportation: driving, bicycling, or walking (travel speeds, in kilometers per hour, are given in table S1; appendix p 5). When calculating travel speeds for bicycling and walking, the slope of the terrain and the direction of travel were taken into account. Detailed methods for constructing the Friction Surface Raster map are given in the SM.

**Figure 2:**
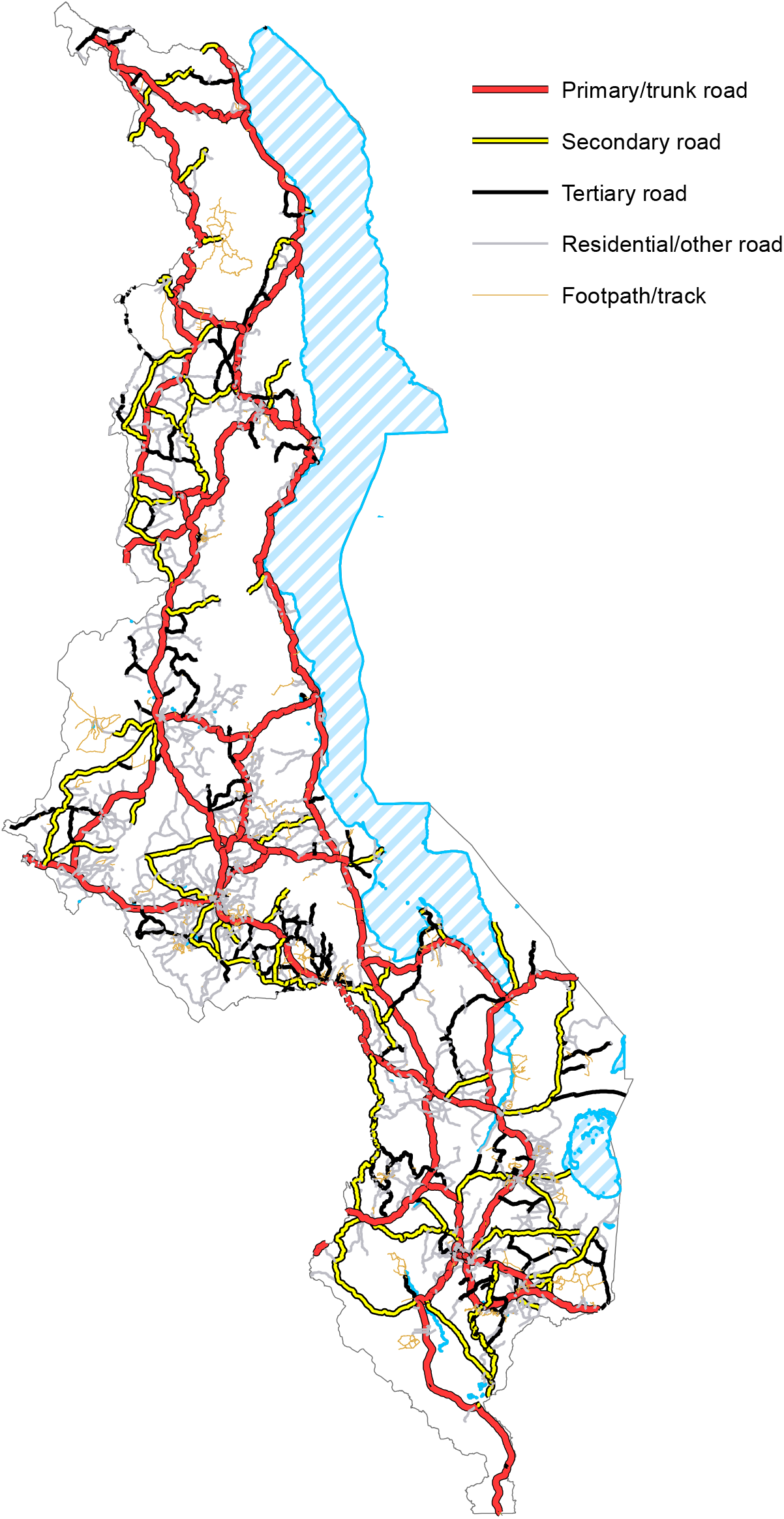
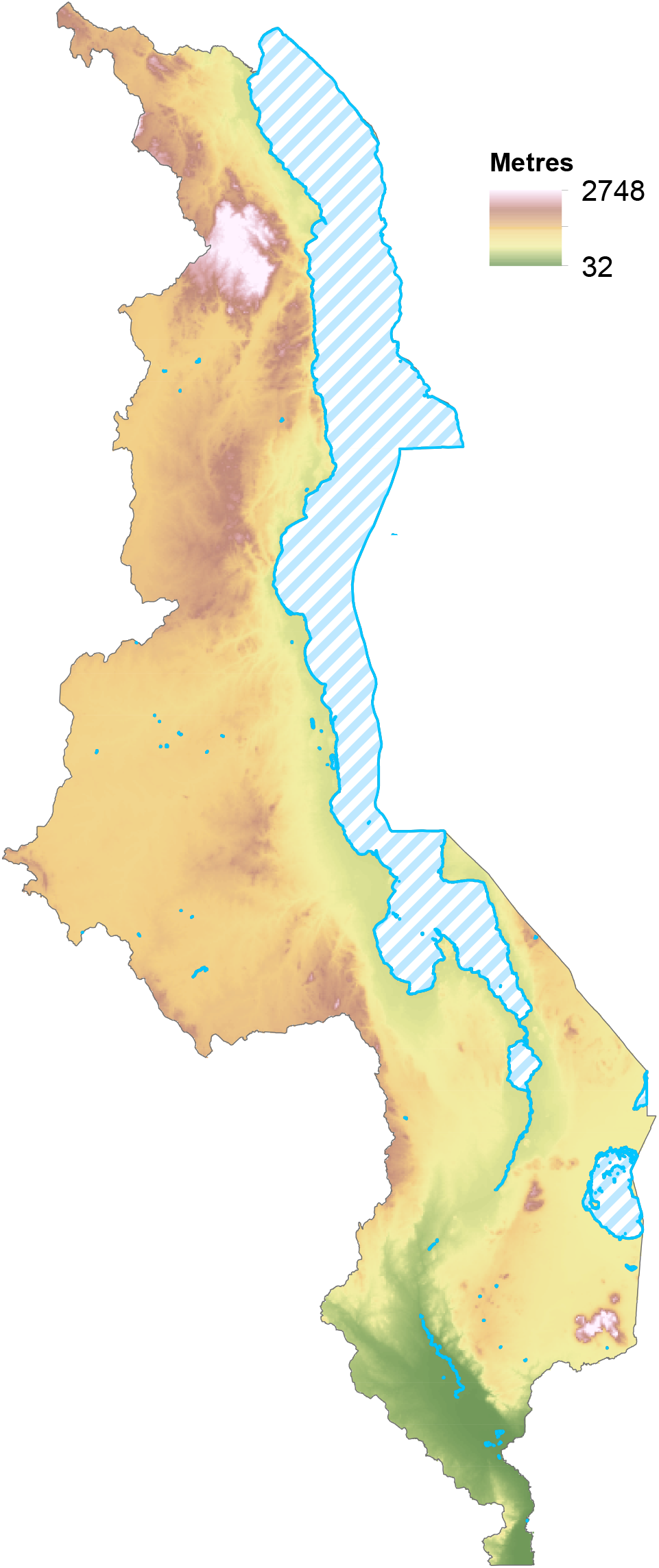
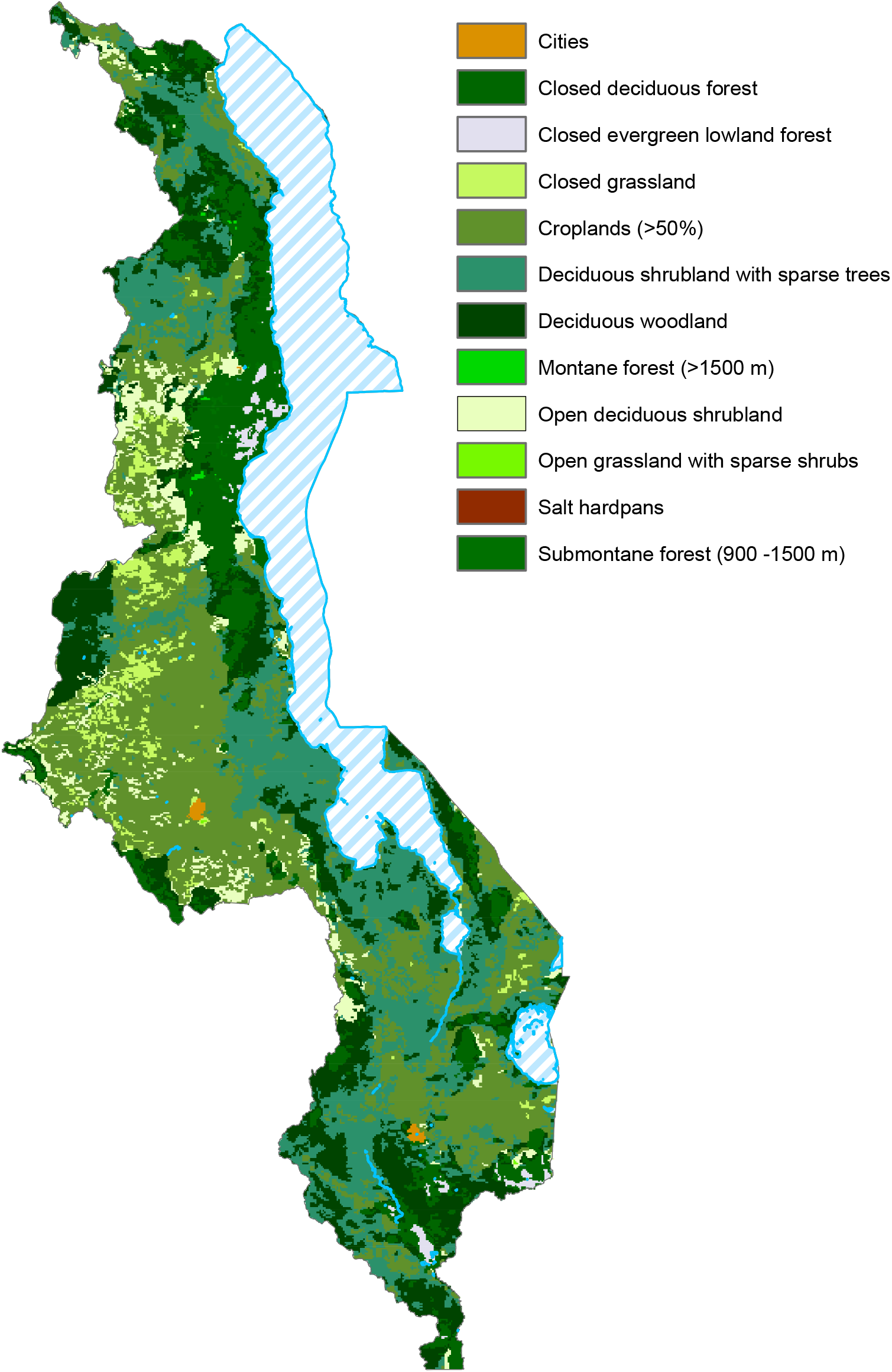
Components of the Friction Surface Raster map. **(A)** Road and water body networks throughout Malawi. **(B)** Topographic map of Malawi. (**C)** Map showing twelve types of landcover.

We used our geospatial modeling framework to determine the effect that the mode of transportation used to travel to HCFs has on the coverage of treatment. To make this determination, we calculated the maximum catchment size of every HCF in Malawi based on a maximum travel-time of one hour and assuming that PLHIV could: (i) drive, bicycle, and walk, (ii) bicycle and walk, or (iii) only walk. The maximum catchment size was calculated by estimating, all around the HCF, the maximum distance that could be covered in a travel-time of one hour; these distances were calculated using AccessMod software.^24^ Additionally, we calculated Maximum Achievable Coverage (MAC) curves; these curves enabled us to estimate the travel-times that would be needed to achieve any specific coverage level, based on a specified mode of transportation.

A MAC curve is a cumulative distribution function: it shows the percentage of PLHIV who can reach their nearest HCF within a specific time. We created the MAC curves by estimating, for each PLHIV in Malawi, their travel-time to the nearest HCF (detailed methods for calculating travel-times are given in the SM), ranking the travel-times from the shortest to the longest, and then creating the cumulative distribution function. The shortest travel-times are for PLHIV who live closest to the HCFs, the longest travel-times are for PLHIV who live furthest from the HCFs.

## Results

The Epidemic Surface Prevalence map reveals that there is substantial geographic variation in the severity of the HIV epidemic throughout the country: prevalence is very high in the cities (~30%), fairly high in fishing villages along Lake Malawi, and very low (2%) in some rural areas (figure 3A). The DoI, which reflects both the geographic variation in population density and the geographic variation in HIV prevalence, varies over three-fold; from >1,000 PLHIV per km^2^ in the cities to less than one PLHIV per km^2^ in rural areas (figure 3B, and figure S3; appendix p 8).

**Figure 3:**
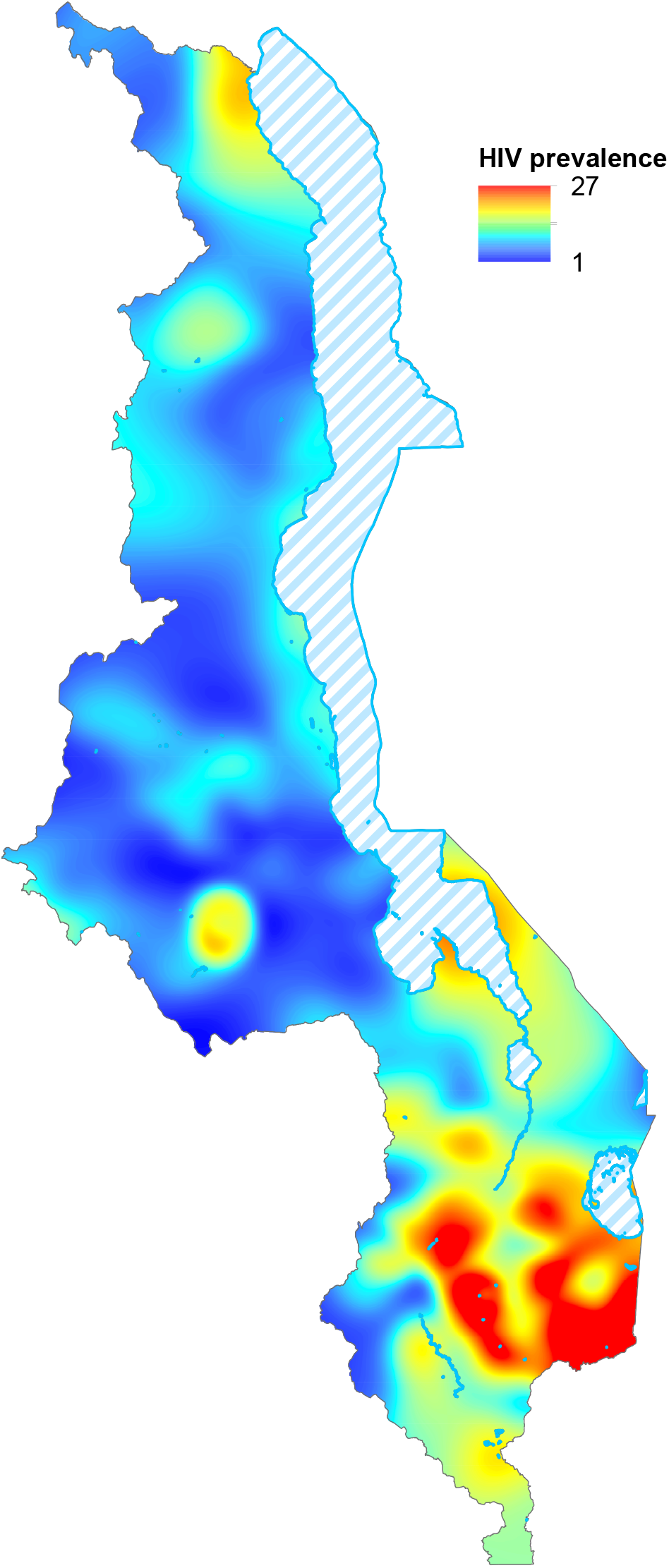
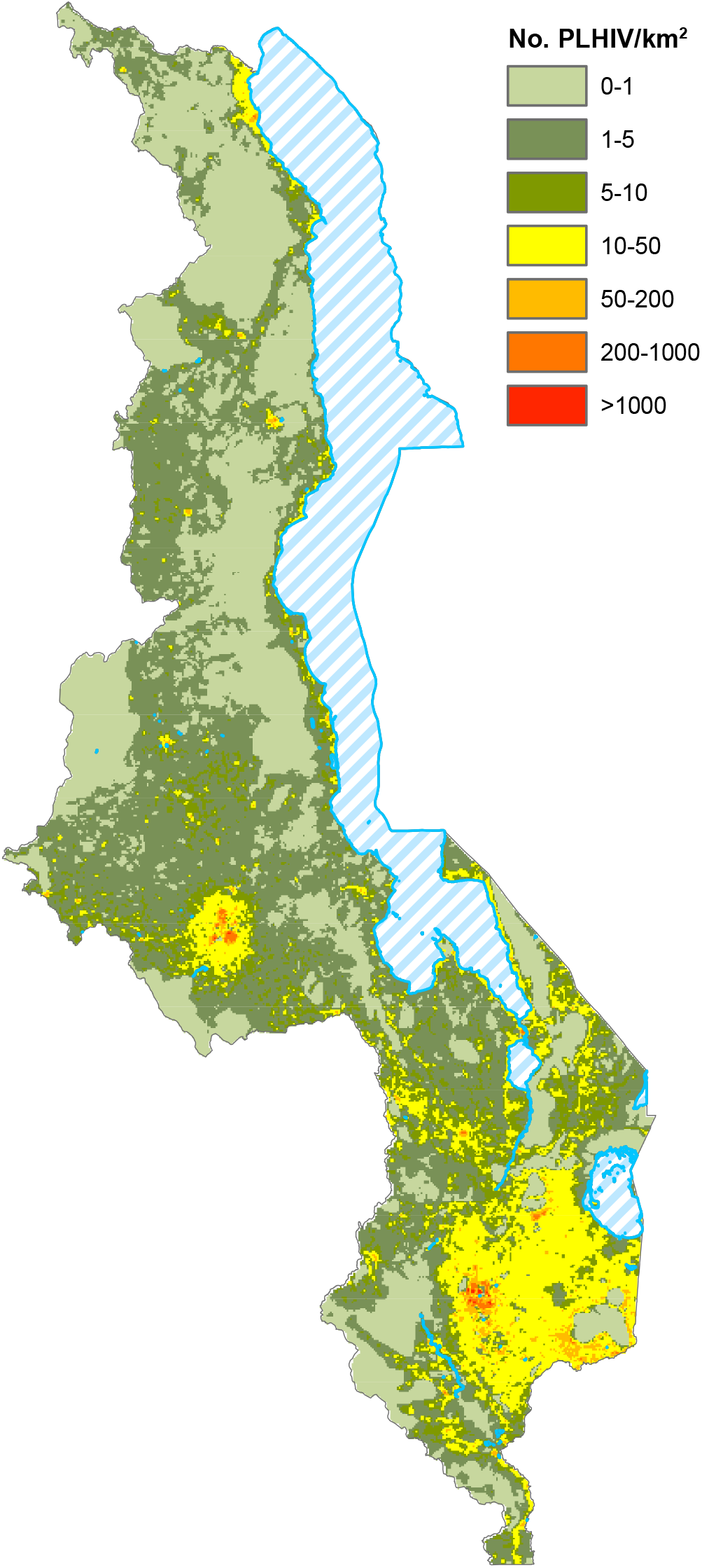
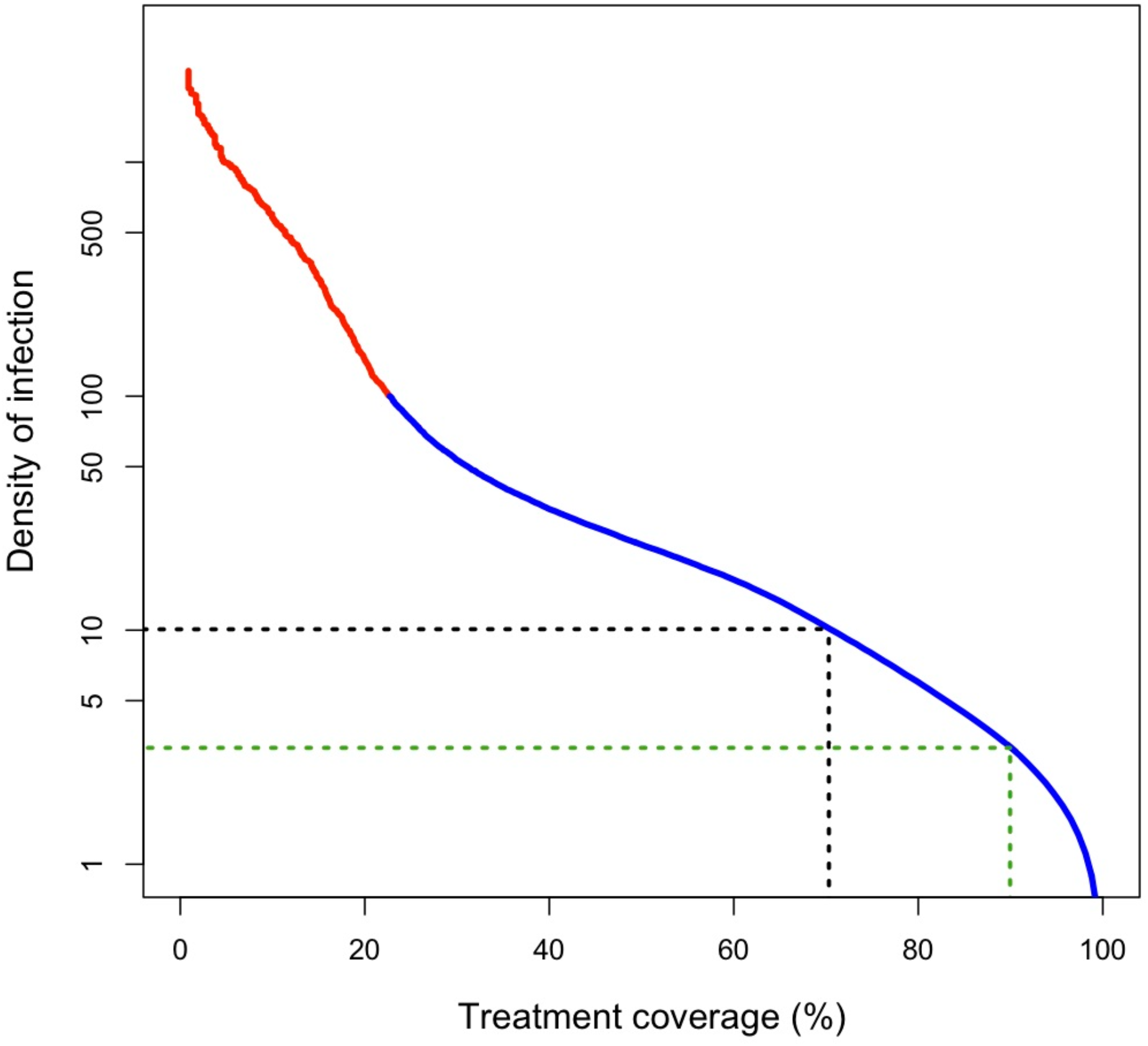
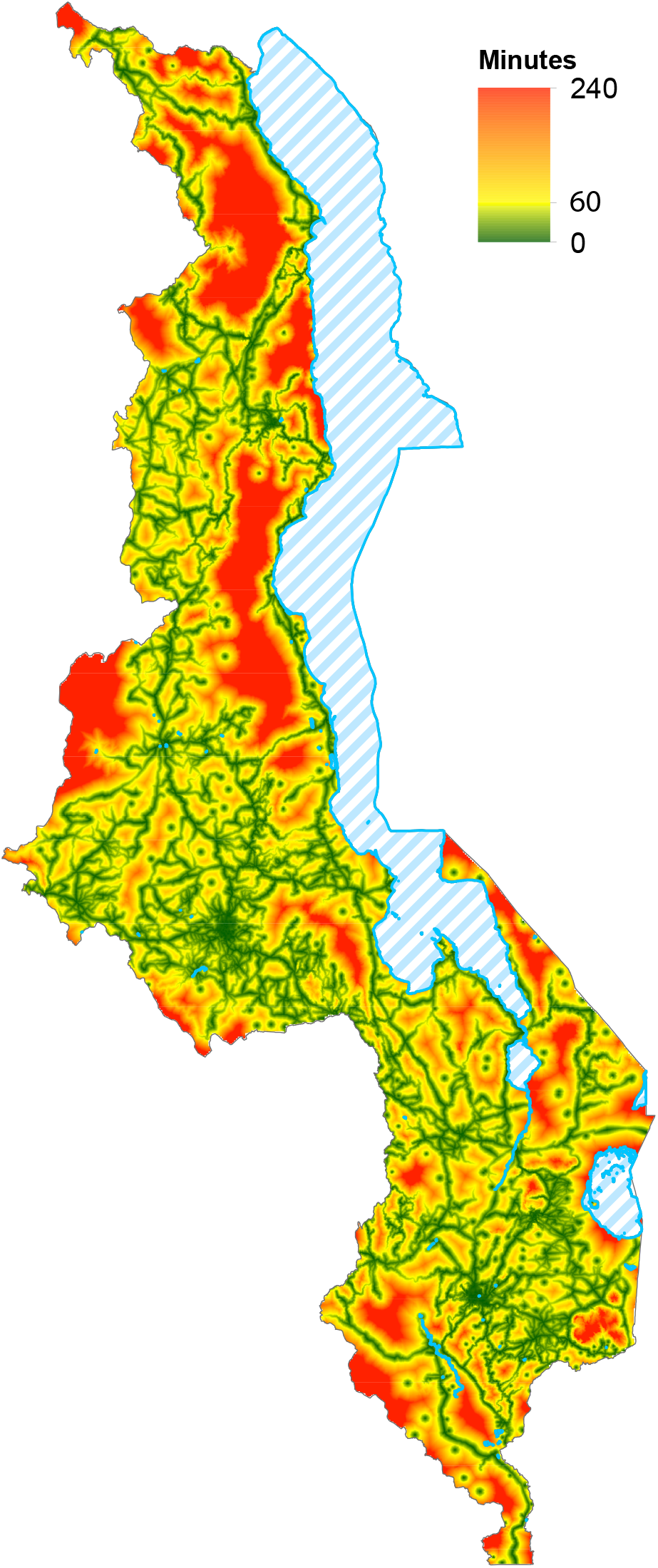
Results on spatial epidemiology and geographic accessibility to HCFs. **(A)** Epidemic Surface Prevalence map showing the prevalence of HIV infection in men and women, aged 15–49 years old. (**B)** DoI map shows the number of PLHIV (15–49 years old) per square kilometer throughout Malawi. (**C)** Epidemic Concentration Curve,^22^ based on data from the DoI map. The Y-axis shows the number of PLHIV per square kilometer, and is plotted on a logarithmic scale. Dotted black line shows DoI at 70% treatment coverage, dotted green line shows DoI at 90% treatment coverage. **(D)** Geographic accessibility to treatment map. The map shows impedance (defined in terms of travel-time) to the nearest HCF from any location in the country.

Figure 3C is a semi-logarithmic plot of the data from the DoI map. This type of plot is known as an Epidemic Concentration Curve;^22^ this curve enables the epidemic to be characterized within a demographic context. It shows the percentage of the epidemic that is concentrated in urbanized areas in Malawi (red data) and the percentage of the epidemic that is dispersed in rural and semi-rural areas (blue data). There is no universal definition of urbanization based on population density. Here, we define - based on the DoI map - an urbanized area as one that has a DoI of greater than 100 PLHIV per square kilometer. Notably, ~80% of PLHIV can be seen to be living in rural and semi-rural areas (figure 3C, blue data). Outside the urbanized areas, the DoI can be seen to drop fairly abruptly, more gradually decrease, and then drop precipitously at a low DoI. The shape of the curve shows that finding - and treating - a high percentage of PLHIV in Malawi, will become progressively more challenging.

Figure 3C also provides important insights into current, and future, treatment programs. Specifically, it can be seen that treatment, at the current coverage level of 70%,^2^ is being provided in some rural areas in Malawi where there is a fairly low DoI: only ~10 PLHIV per km^2^. The plot also shows that to expand treatment programs to reach UNAIDS’ 90% coverage goal will require providing treatment in even more rural areas where there are only ~4 PLHIV per km^2^.

The Friction Surface Raster map (figure S4; appendix p 9) shows that there is substantial geographic variation in impedance, both due to the terrain and the mode of transportation. We estimate that it takes from ~60 seconds (driving on main roads) to ~60 minutes (walking in mountainous areas) to cross an area of one km^2^ in Malawi. Using the Friction Surface Raster map, we created a geographic accessibility to treatment map that shows the travel-time to the nearest HCF from anywhere in Malawi (figure 3D); detailed methods for constructing the geographic accessibility to treatment map are given in the SM. The map shows that, due to geographic variations in impedance and the spatial distribution of HCFs, there is substantial urban-rural inequity in geographic accessibility to treatment; the most urbanized areas are in the South and the most rural in the North. It takes less than one hour in urban/semi-urban areas to reach a HCF, one to two hours in many rural areas, and up to four hours in remote rural areas.

Figure 4 shows the maximum catchment size of every HCF in Malawi based on a maximum travel-time of one hour and assuming that PLHIV can drive, bicycle, and walk (figure 4A), bicycle and walk (figure 4B), or only walk (figure 4C). The catchment sizes for each HCF are shown by the green data. Notably, there is a substantial increase in catchment size in the predominantly rural areas if PLHIV can bicycle rather than walk: compare figure 4B with figure 4C. As catchment size increases, a greater number of PLHIV live within the catchment area, hence coverage can increase. At the national level, given a maximum travel-time of one hour, the MAC would be ~80% (if all three modes of transportation were available, figure 4A), ~65% (if only bicycles were available, figure 4B), but only ~50% if all PLHIV had to walk (figure 4C). Taken together, these results show that the lack of transportation in rural areas in Malawi are major barriers to achieving very high coverage levels. Additionally, they show that increasing the availability of bicycles in rural areas has the potential to substantially increase treatment coverage at the national level.

**Figure 4:**
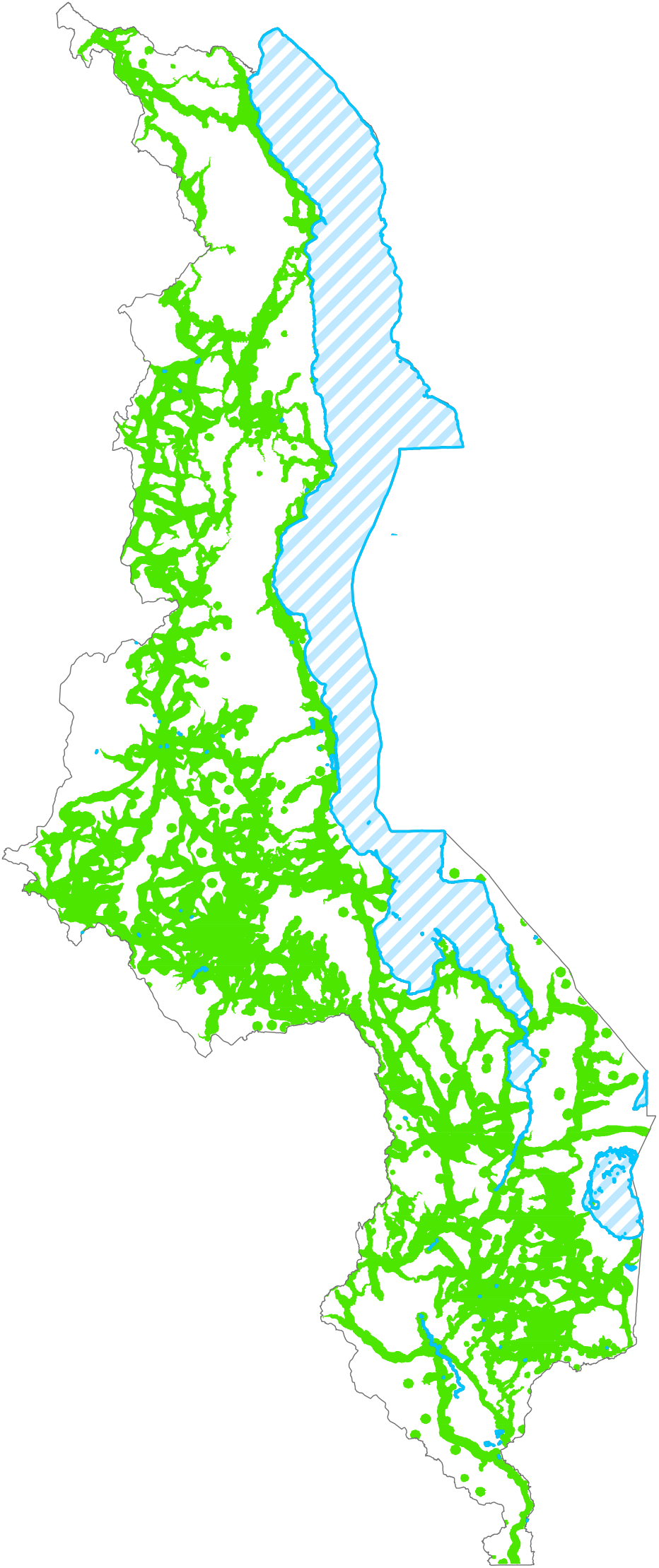
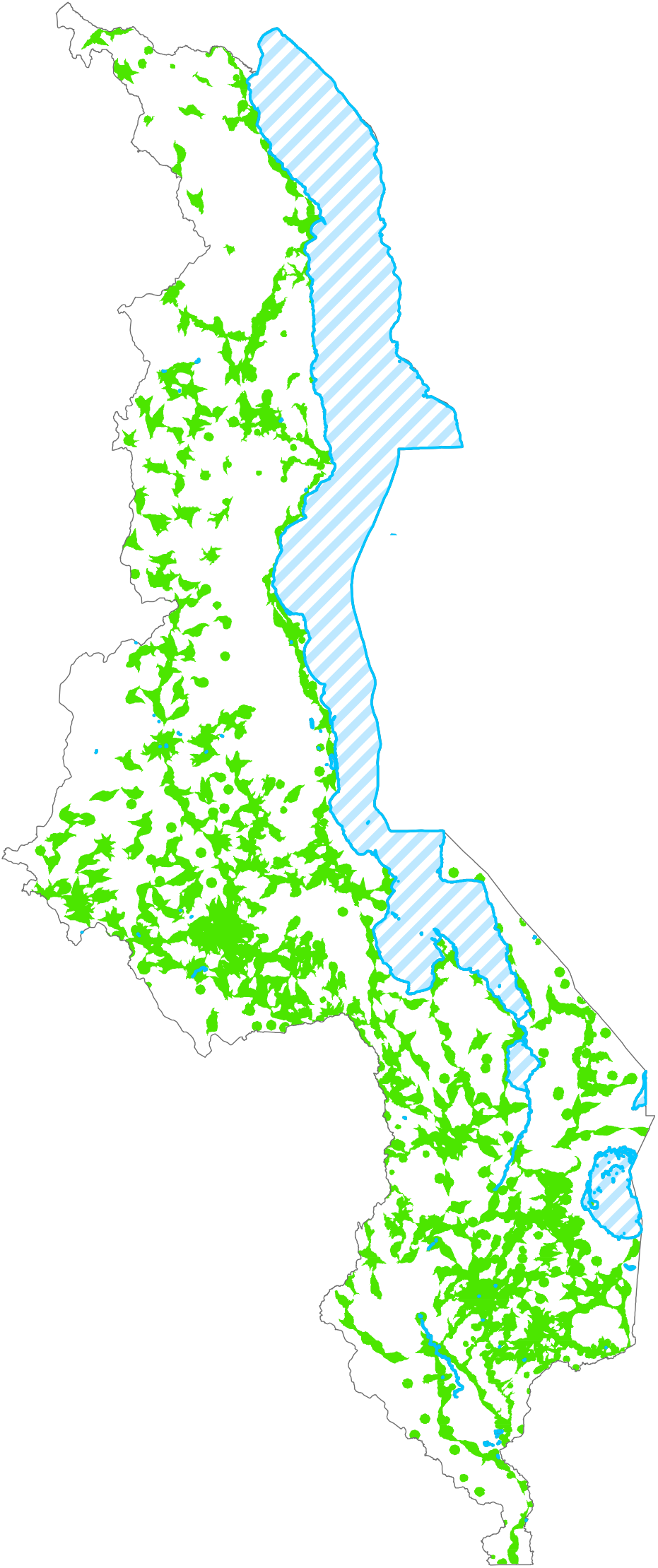
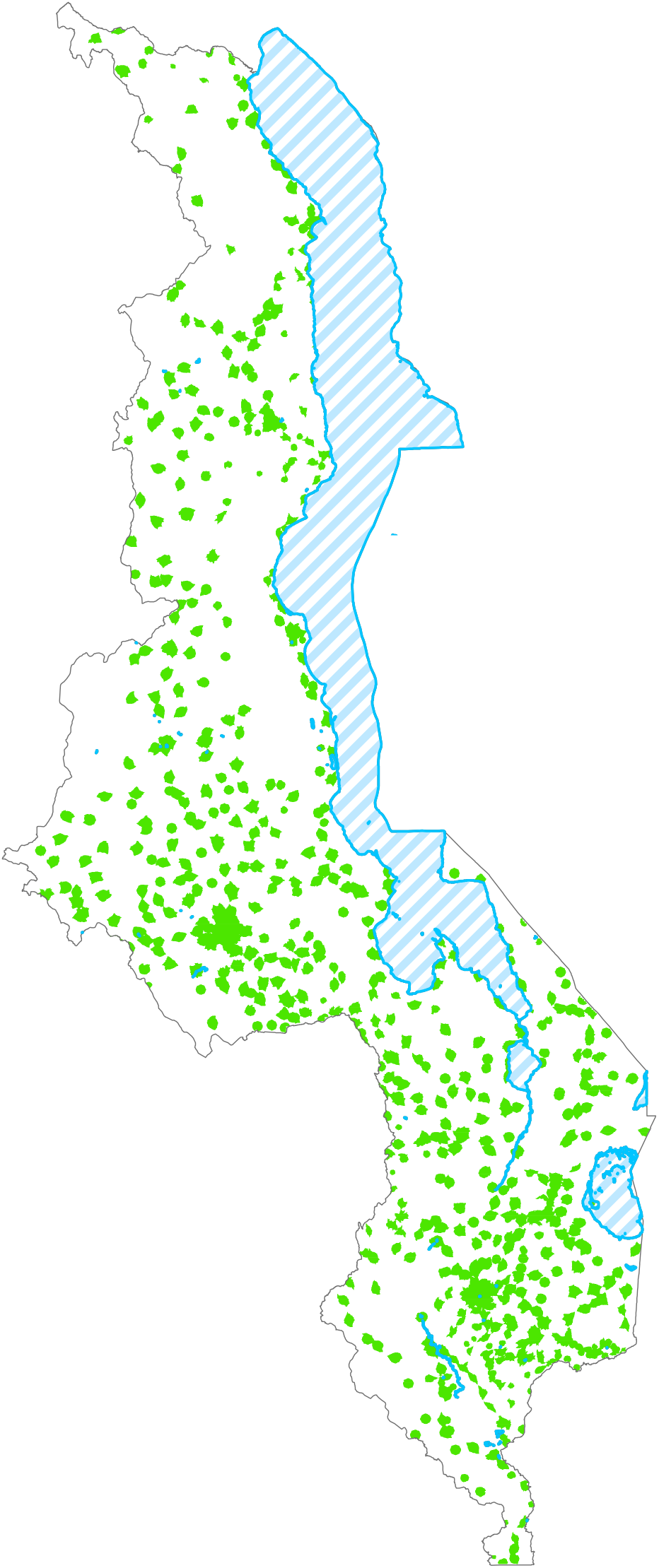
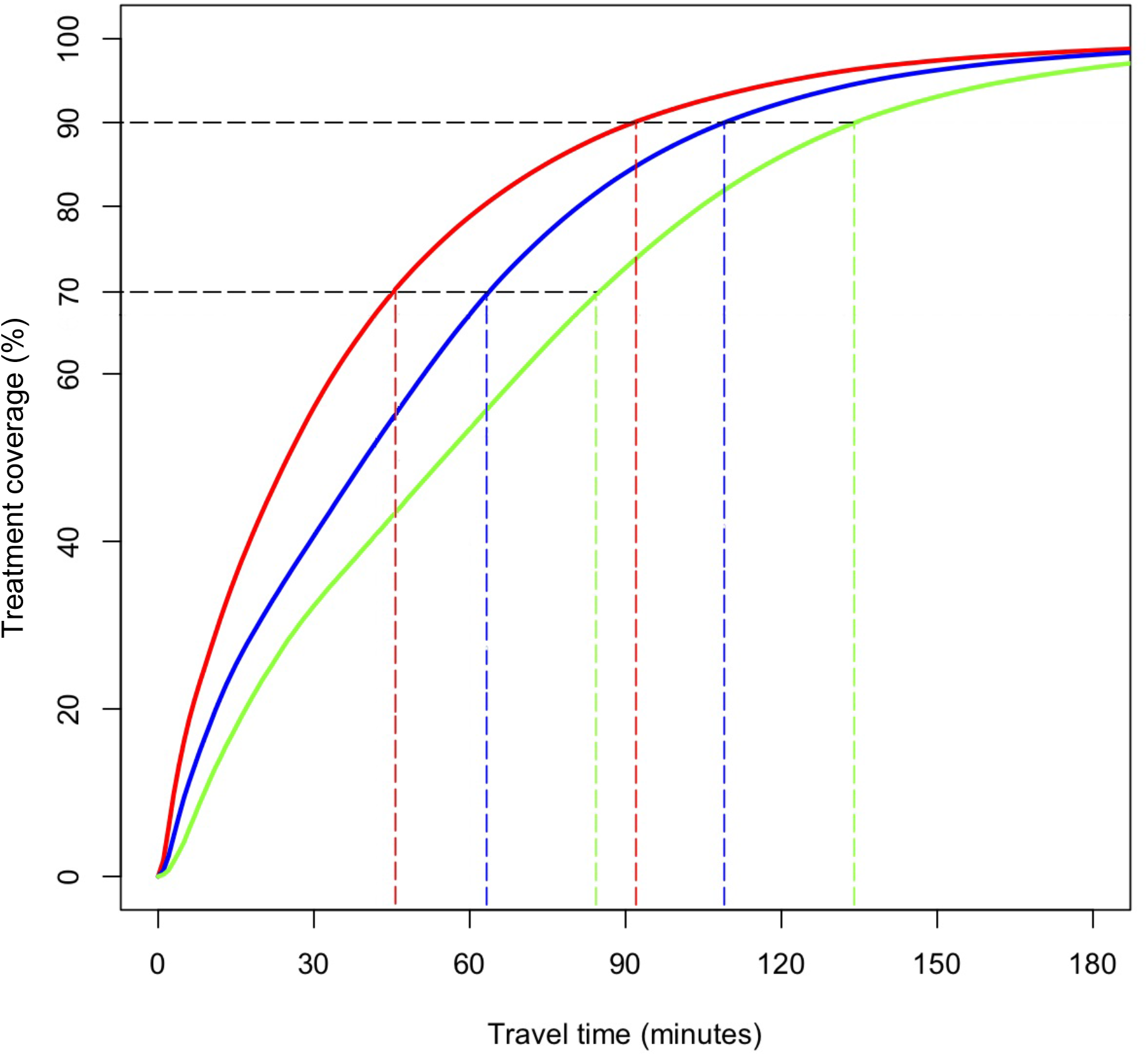
Accessibility by mode of transportation, and MAC Curves. **(A)** Areas (shown in green) where access to a HCF takes less than 60 minutes. Catchment size around each HCF is calculated assuming that individuals can drive, bicycle, and walk. **(B)** Areas (shown in green) where access to a HCF takes less than 60 minutes. Catchment size around each HCF is calculated assuming that individuals can only bicycle and walk. **(C)** Areas (shown in green) where access to a HCF takes less than 60 minutes. Catchment size around each HCF is calculated assuming that individuals can only walk. **(D)** MAC Curves show the percentage of PLHIV who can access a HCF within a given travel-time. Travel-time is defined as the one way travel-time to reach the nearest HCF. The red curve shows the MAC under the assumption that individuals can drive, bicycle and walk; the blue curve shows the MAC under the assumption that individuals can bicycle and walk; the green curve shows the MAC under the assumption that individuals can only walk.

The MAC Curves (figure 4D) show the estimated travel-times to the nearest HCF for all PLHIV in Malawi, both diagnosed and undiagnosed. The three curves show the MAC - at the national level - as a function of the amount of time needed to spend traveling to HCFs and the mode of transportation: walking (green), bicycling and walking (blue), driving, bicycling, and walking (red). Results show that travel-times - at all coverage levels - would be substantially reduced, in comparison with walking, if bicycles were readily available, and used, in rural communities. Notably, the three MAC curves converge at ~95% coverage; this is because, at this coverage level, the remaining HCFs are only accessible by walking. The MAC curves show that, at the current coverage of ~70%,^2^ PLHIV who drive to HCFs can reach them within ~45 minutes (red data), those who bicycle can reach them within ~65 minutes (blue data), and those who walk can reach them within ~85 minutes (green data).

Our results show that increasing coverage above 70% will become progressively more challenging, this is due to the non-linearity of the relationship between coverage and the travel-time to HCFs (figure 4D). Our results show that it will only be possible to achieve UNAIDS’ 90% coverage goal in Malawi if many of the PLHIV (who have yet to initiate treatment) are willing, and able, to spend a great deal of time travelling to access treatment. Notably, many of these individuals will need to spend almost twice as much time traveling to reach a HCF as those who are already on treatment. Those with access to motorized transportation will need to travel for up to an hour and a half to reach the nearest HCF (red data), those with bikes will need to travel for up to ~1·8 hours (blue data), and those who walk will need to travel for up to ~2·2 hours (green data). Therefore, in the absence of bicycles, many PLHIV living in rural areas will need to walk for approximately four and a half hours round-trip to access treatment: increasing the availability of bicycles would reduce the round-trip travel-times by approximately one hour. Notably, the longer it takes a PLHIV to reach a HCF, the less likely they are to access and/or adhere to treatment.^6^

## Discussion

Our study had two objectives. First, to evaluate the potential impact of the necessity of traveling long distances to access HIV treatment and the lack of transportation in rural areas as barriers to eliminating HIV in Malawi. Specifically, their impact on preventing treatment coverage from reaching UNAIDS’ 2030 target of 90%. We found that although HIV prevalence is much higher in urban areas than in rural areas, the vast majority of PLHIV live in rural areas; many live in settlements where there is a very low density of infection. We also found that there is a nonlinear relationship between travel-time to access treatment and the attainable coverage level. As a consequence of this relationship, increasing coverage levels above the current fairly high level of ~70% will become progressively more difficult. Taken together, our results show that geographic inaccessibility to HIV treatment coupled with the lack of transportation in rural areas are substantial barriers to HIV elimination. Our second objective was to evaluate the potential impact of a novel intervention strategy for improving treatment coverage: increasing the availability of bicycles in rural areas. We found that this intervention could substantially increase treatment coverage levels in rural areas, and has the potential to make it possible for Malawi to reach UNAIDS’ 90% elimination goal. Notably, the geospatial modeling framework that we have developed can be used in multiple ways as a health policy tool for designing and evaluating strategies for controlling HIV in other countries in SSA.

Improving geographic accessibility to HIV treatment will have many additional healthcare benefits in Malawi,^20^ as well as increasing the life expectancy for many PLHIV. For example, it will increase the proportion of HIV-infected women who are on treatment before getting pregnant; this will reduce mother-to-child-transmission, as well as improve antenatal care for all pregnant women. Increasing geographic accessibility to healthcare also has the potential to prevent a substantial number of deaths in children under five years of age. Many of these deaths in Malawi - as in many other countries in SSA - are due to a small number of common causes (diarrhea, malaria, and poor prenatal care); these are preventable through services that are often available at local clinics. Improving access is also likely to increase the coverage of treatment rates for tuberculosis and immunization rates for childhood diseases.

When estimating travel-times to access treatment we have taken into account settlement patterns, population density, the spatial diffusion of the HIV epidemic, road and water body networks, landscape features, topography, the type of transportation that is available, and the number and geographic distribution of HCFs. We did not include the effects of weather or road conditions that could increase the difficulty of travel, or that some clinics have a limited capacity to treat a large number of patients. Furthermore, our estimates are based on walking speeds for an average healthy individual; PLHIV who are ill, pregnant and/or travelling with children are likely to walk substantially slower than average and take longer to reach HCFs. Even if PLHIV travel to HCFs, treatment may not always be available due to drug stock-outs.^8,10,12^ Our modeling framework can be expanded to take all of these factors into consideration. Notably, including any of these factors will make travel-times substantially longer than those estimated for this analysis; therefore, our current estimates should be considered optimistic, and could potentially be under-estimates.

We have considered two major barriers to reaching UNAIDS’ 90% coverage goal: distance to healthcare and limited transportation. However, there are additional barriers that prevent PLHIV from accessing healthcare,^8,26-28^ e.g., stigma, discrimination, fear of disclosure, depression, fear of drug side effects, transportation costs, the need to take time off work, fear of violence, the availability of HIV-testing and counseling and (for some women) needing to obtain permission to travel.^29,30^ Creative solutions will need to be found to overcome the multiple barriers that prevent PLHIV accessing - and adhering to - treatment.^7,31,32^ Here we have proposed one potential solution to increasing geographic accessibility to healthcare in a UNAIDS designated Fast-Track country, Malawi. Many other solutions are possible to increase geographic accessibility to care, e.g., further decentralization of care, improved transportation, mobile clinics, drones for delivering medication, community medication groups (i.e., only one member of the community visits the clinics and collects medications for all PLHIV in the community) and differentiated care initiatives. A single solution will not solve the problem; a combination of solutions is needed.

Our results have significant implications for HIV policy in Malawi. Our results indicate that it is unlikely that treatment coverage will have reached UNAIDS target of 81% for 2020, and that it is extremely unlikely - based on the current healthcare infrastructure - that coverage will reach UNAIDS 2030 target of 90% coverage. The current policy in Malawi is to focus on HIV-testing for key and vulnerable populations in high-yield settings. This policy is driven by the fact that HIV prevalence and the incidence rates are high in these populations. However, as we have found, the vast majority of PLHIV live in rural areas - some of them in extremely remote areas: many of these PLHIV may not have been diagnosed. We suggest that individuals who live in very rural settings should also be considered as a key population even though HIV prevalence (and therefore incidence) is low. Focusing HIV-testing in very rural areas, due to the low density of infection, will result in low yields; i.e., a low percentage of HIV-infected individuals per number tested. Therefore this strategy will not be efficient or cost-effective; however - based on our results - it will be essential to employ this strategy in order to reach UNAIDS’ coverage goals for 2030.

Our results demonstrate that there is a need for international health policy organizations (such as the WHO, UNAIDS, and PEPFAR), as well as Governments of HIV-afflicted countries in SSA, to evaluate the feasibility of the policies - and the goals - that they propose. As we have shown there are severe constraints that are imposed by demography (e.g., settlement dispersal and population density), landscape features, and the transportation infrastructure that can limit a country’s ability to achieve desired goals. We recommend that these constraints should be included in the epidemiological and economic models that are used as health policy tools to evaluate the cost-effectiveness, and to predict the impact, of HIV interventions and treatment programs. Furthermore, Governments in SSA often determine access to healthcare based on distance. We suggest that a more appropriate and equitable measure than distance to HCFs would be travel-time to HCFs.

We have proposed and evaluated a novel intervention for helping in the control of HIV epidemics: increasing the availability of bicycles in rural areas. Our results have shown that this intervention has the potential to substantially increase coverage levels, to make the 90% coverage goal attainable, and hence could help eliminate HIV in Malawi. The Government of Malawi is unlikely to implement this intervention. However, we believe that - if second-hand bicycles are used - this intervention is feasible, and could be funded through crowd-sourcing. Donated second-hand bicycles are already being distributed, on a small scale, by several charities in Malawi. These charities have multiple objectives, improving healthcare, education, economic development, and gender equality.^33,34^ Therefore, we suggest that expanding their efforts in increasing accessibility to bicycles - as well as increasing access to healthcare - will also increase educational opportunities, help economic development in rural areas, and help address the problems of gender inequality.

Our study has several limitations. To make our estimates of travel-time we have assumed that PLHIV travel to their nearest HCD. People do not always attend their nearest clinic.^35^ However, when there are long distances to travel to access healthcare, many people are likely to attend their nearest HCF. If people do not attend their nearest clinic, travel-times will be even longer than we have estimated, and achieving the 90% coverage level will be even more unlikely. We have evaluated the time it takes to travel to HCFs, but we have not presented any data that suggests people living further from care are actually less likely to receive care in Malawi. However, two recent studies^36,37^ have shown that such a connection exists: decreased travel distance is associated with increased treatment initiation and retention. One study, based in the Neno District in Malawi, found that patients living more than 8 kilometers away from their HCF had a greater hazard (adjusted Hazard Ratio: 1·68, 95% Confidence Interval 1·49–1·89) of being lost to follow-up than patients living within 8 kilometers of their HCF; this effect was independent of age and gender.^36^ The second study took place in rural KwaZulu-Natal in South Africa: the likelihood of accessing treatment decreased by 27% with every square-root transformed kilometer from the nearest HCF.^37^

Our results have significant implications for HIV policy in other HIV-afflicted countries in SSA. Malawi is one of UNAIDS’ designated Fast-Track countries.^12^ Distance to healthcare and lack of transportation are major barriers to accessing HIV treatment throughout SSA. Our study is the first that shows they can be substantial barriers to HIV elimination. Notably, the 21 other designated Fast-Track countries in SSA have similar characteristics to Malawi: they all have predominantly rural populations, severe generalized HIV epidemics,^38^ and limited healthcare infrastructure. The geospatial modeling framework that we have developed can be used to analyze geographic accessibility to healthcare in any of the Fast-Track countries, and in at least ten other countries in SSA; the country-specific data needed to conduct these analyses are publicly available. We believe that it is essential to conduct similar analyses for these other countries, to those that we have conducted for Malawi, in order to quantify the effect of distance to healthcare and transportation on preventing HIV elimination in SSA.

## Data Availability

All data associated with the study are available within the manuscript or from the authors upon reasonable request.

## Acknowledgements

We are grateful to Eugenio Valdano and Nelson Freimer for discussions throughout the course of this research.

## Contributors’ statement

SB developed the concept, interpreted results, and drafted the manuscript. LP programmed the model, generated figures, and interpreted results. JTO and LD interpreted results and contributed to writing. All authors read and approved the final manuscript.

